# COVID-19 Genome Surveillance: A Geographical Landscape and Mutational Mapping of SARS-CoV-2 Variants in Central India over Two Years

**DOI:** 10.1101/2023.03.22.23287566

**Authors:** Krishna Khairnar, Siddharth Singh Tomar

## Abstract

Reading the viral genome through whole genome sequencing enables the detection of changes in the viral genome. The rapid changes in the SARS-CoV-2 viral genome may cause immune escape leading to an increase in the pathogenicity or infectivity. Monitoring mutations through genomic surveillance helps understand the amino acid changes resulting from the mutation. These amino acid changes, especially in the spike glycoprotein, may have implications on the pathogenicity of the virus by rendering it immune-escape. The region of Vidarbha in Maharashtra represents 31.6% of the total area and 21.3% of the total population of the state. In total, 7457 SARS-CoV-2 positive samples belonging to 16 Indian States were included in the study, out of which 3002 samples passed the sequencing quality control criteria. The metadata of 7457 SARS-CoV-2 positive samples included in the study was sourced from the Integrated Health Information Platform. The metadata of 3002 sequenced samples, including the FASTA sequence, was submitted to the Global initiative on sharing Avian Influenza Data and the Indian biological data centre. This study identified 104 different SARS-CoV-2 pango-lineages classified into 19 clades. We have also analysed the mutation profiles of the variants found in the study, which showed eight mutations of interest, including L18F, K417N, K417T, L452R, S477N, N501Y, P681H, P681R, and mutation of concern E484K in the spike glycoprotein region. The study was from November 2020 to December 2022, making this study the most comprehensive genomic surveillance of SARS-CoV-2 conducted for the region.

## 1. INTRODUCTION

Viruses continue to mutate as they spread within the host population, develop into variants, and deviate genomically from the original virus. It is important to understand these changes in the viral genome and how they impact the aetiology of the virus. It is required to read the viral genome through whole genome sequencing (WGS) to detect changes in the viral genome. As the viral genome is comparatively smaller than the prokaryotic or eukaryotic genome, the frequency of mutations in the viral genome is comparatively high. The SARS-CoV-2 virus originated in Decemebr’2019 as a wild type, further diversifying into several variants of SARS-CoV-2. The rapid changes in the SARS-CoV-2 viral genome may cause immune escape leading to an increase in the pathogenicity or infectivity [1]. Monitoring mutations through genomic surveillance allows the researchers to understand the amino acid changes resulting from the mutation. These amino acid changes, especially in the spike glycoprotein, may have implications for the pathogenicity of the virus by making it escape the monoclonal antibody therapies and vaccine-induced immunity [2]. Hence, it is important to keep track of the changes in the viral genome to devise effective mitigation strategies. To track the emerging variants of SARS-CoV-2, healthcare authorities across the globe initiated genome surveillance programs. The government of India also initiated a nationwide SARS-CoV-2 genome surveillance network known as the Indian SARS-CoV-2 Consortium on Genomics (INSACOG), a forum established on 30 December 2020 under the Ministry of Health and Family Welfare to keep track of circulating SARS-CoV-2 variants in India [3].

This region of Vidarbha in Maharashtra represents 31.6% of the state’s total area and holds 21.3% of the total population. It is also worth mentioning that the state of Maharashtra has emerged as a hotspot of SARS-CoV-2 infections since the pandemic’s beginning. The origin of the virulent delta variant that drove the second wave of SARS-CoV-2 in India and around the globe was the district of Amravati in the Vidarbha region of Maharashtra [4,5]. The largest city of the Vidarbha region is Nagpur, situated strategically at the centre of the north-south axis of India; Nagpur, an important urban centre of the region, also serves as a medical hub for the population living in surrounding rural and tribal areas. Nagpur is the third largest city in the Indian state of Maharashtra and the fourteenth largest city in India by population [6]. Any outbreak in Nagpur city could easily spread to other regions of India. Therefore, the findings of genome surveillance from the Vidarbha region hold great significance for understanding the dynamics of SARS-CoV-2 in India to control the spread of the SARS-CoV-2 virus in the population. This study was part of national SARS-CoV-2 genome surveillance efforts for the central region of India.

Keeping this in view, the Council of Scientific and Industrial Research-National Environmental Engineering Research Institute (CSIR-NEERI) Nagpur started the genomic surveillance of SARS-CoV-2 primarily for the Nagpur district of Maharashtra as gargle-based genome surveillance [7]. After becoming part of INSACOG, the genome surveillance mandate expanded to other districts of Maharashtra and other states of India like Tamil Nadu and the Union Territory of Jammu & Kashmir. In total, 7457 SARS-CoV-2 positive samples belonging to 16 Indian States were included in the study, out of which 3002 samples passed the sequencing quality control criteria **(Figure 1)**. The duration of this study was from November 2020 to December 2022, which makes this study the most comprehensive genomic surveillance of SARS-CoV-2 conducted for the region.

**Figure 1:**
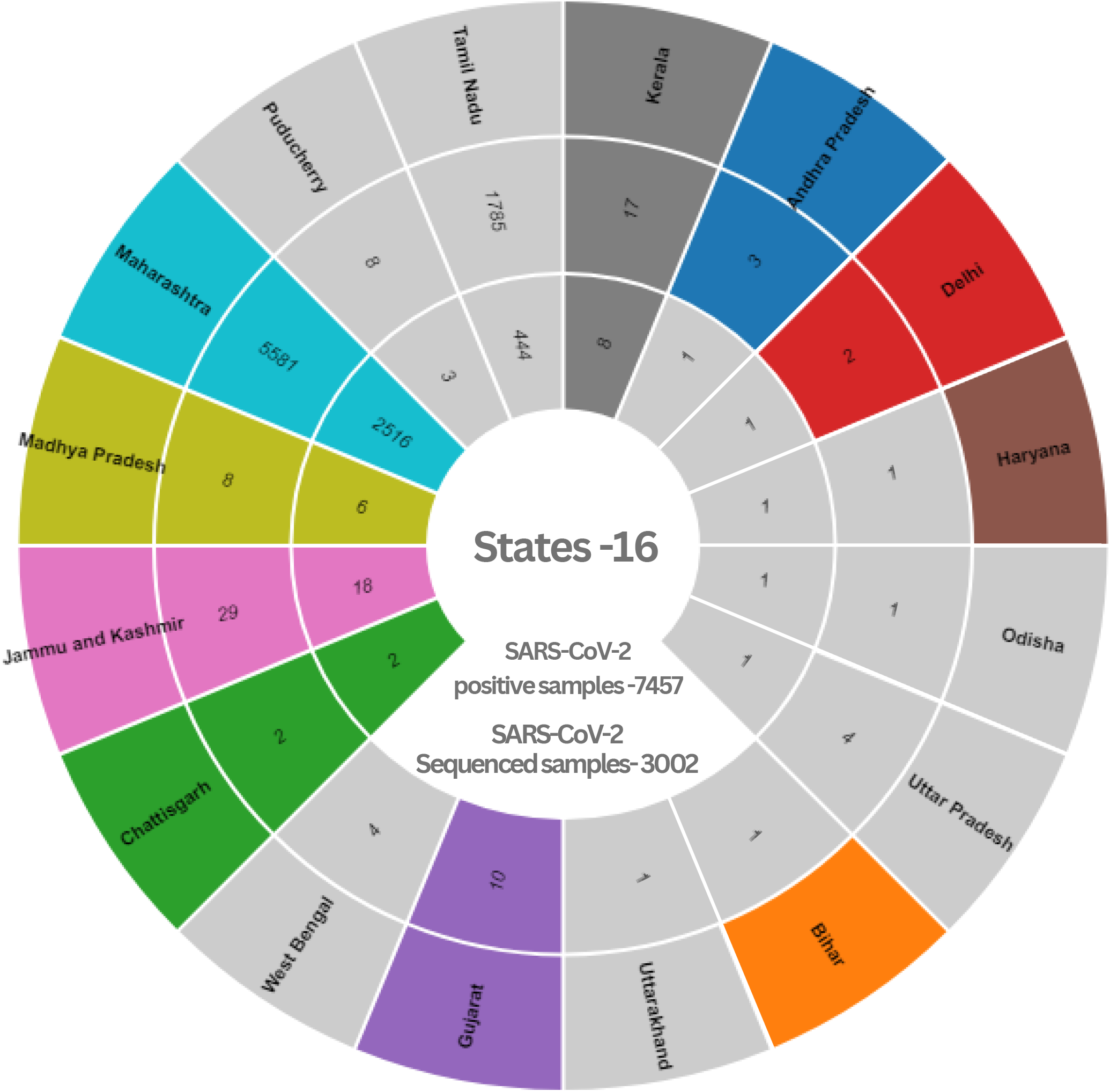
Summary of the study’s SARS-CoV-2 samples and state-wise distribution.

## 2. MATERIALS AND METHOD

### 2.1 Data collection

The metadata of 7457 SARS-CoV-2 positive samples included in the study was sourced from the Integrated Health Information Platform (IHIP) [8]. The hospitalisation and vaccination status information was obtained from the Indian council of medical research (ICMR) COVID-19 data portal [9]. Within the vaccine information available group, 27.04% of patients were vaccinated with Covaxin, 72.13% were vaccinated with the Covishield vaccine, and 0.8% with the Sputnik V vaccine. In the study population of 7457, the overall gender distribution was 52.9% (3950) male, 47.02% (3506) female and 0.01% (1) transgender. In this study, the percentage distribution of the cases within the age groups was 0.36% (0 to <2 years), 0.63% (2 to <5 years), 5.8% (5 to <15 years), 65.7% (15 to <50 years), 18.5% (50 to <65 years), and 8.97% (≥ 65 years). The age distribution was according to the WHO Global Epidemiological Surveillance Standards for Influenza [10].

### 2.2 Materials

The SARS-CoV-2 viral RNA extraction was performed for gargle-based genome surveillance using an RNA release buffer containing Tris-EDTA and Proteinase K. The SARS-CoV-2 viral RNA extraction for OPS-NPS-based genome surveillance was performed using Insta NX automated nucleic acid extractor. Applied Biosystems QuantStudio 3 and StepOnePlus qRT-PCR machines were used for qRT-PCR, cDNA synthesis, PCR tiling, and rapid barcoding. Qubit 4 Fluorometer by Thermo Fisher Scientific was used to quantify nucleic acids for quality control (QC) of DNA libraries for Oxford Nanopore Technology (ONT)-based WGS. SARS-CoV-2 WGS was done on an Mk1C 6.3.9 and Mk1B ONT MinION sequencing platform.

### 2.3 Sample collection and RNA extraction

Out of 7457 SARS-CoV-2 positive samples included in the study, 707 were collected by saline gargle collection, and 6750 samples were collected using OPS-NPS VTM swabs from November 2020 to December 2022. The SARS-CoV-2 samples were collected using a saline gargle, and one-step RNA isolation from gargle samples was performed per the method developed by Khairnar et al. [7]. A video documentary of SOP on CSIR-NEERI’s saline gargle-based SARS-CoV-2 molecular testing is available [11]. The SARS-CoV-2 samples collected using a nasopharyngeal-oropharyngeal (NP/OP) swab were suspended in a viral transport medium (VTM). The RNA from VTM swab samples was extracted using an automated RNA extractor using MBIN013 Insta NX Viral RNA Purification Kit according to the manufacturer’s protocol (HIMEDIA) [12]. The isolated RNA samples were used immediately or stored at -80^°^C till further use.

### 2.4 RT-PCR for detecting SARS-CoV-2

qRT-PCR was performed to quantitatively detect SARS-CoV-2 in samples using MBPCR255 Hi-PCR COVID-19 Triplex Probe PCR Kit (HIMEDIA) [13] and/or Meril SARS-CoV-2 kit [14] The primer-probe sets in the kits specifically detect viral RNA from SARS-CoV-2. The samples with a Ct value of ≤38 for SARS-CoV-2 genes were considered positive for SARS-CoV-2 by RT-PCR.

### 2.5 SARS-CoV-2 whole genome sequencing

Out of 7457 samples, only 3002 samples qualified for SARS-CoV-2 WGS according to the Ct value selection criteria of GISRS [15 Refer to the criteria document of GISRS, which we had used in the review paper]. The cDNA synthesis was done using the TAKARA Prime Script RT reagent kit (RR037A) [16]. The sequencing libraries were prepared by multiplex PCR tiling according to the protocol of ONT [17]. The prepared cDNA libraries were sequenced using MinION Mk1C or MinION Mk1B.

### 2.6 Viral load dynamics in SARS-CoV-2 samples

The sample’s cycle threshold (Ct) value in RT-PCR exhibits inverse relation with the viral load. The study with OPS-NPS-based genome surveillance showed sequencing success, with samples exhibiting relatively lower Ct values. However, the saline gargle-based genome surveillance study showed sequencing success even with samples exhibiting relatively higher Ct values. Gargle-based genome surveillance enabled SARS-CoV-2 WGS of positive samples even with a lower viral load, exhibiting Ct values as high as >35 (**Figure 2)**.

**Figure 2:**
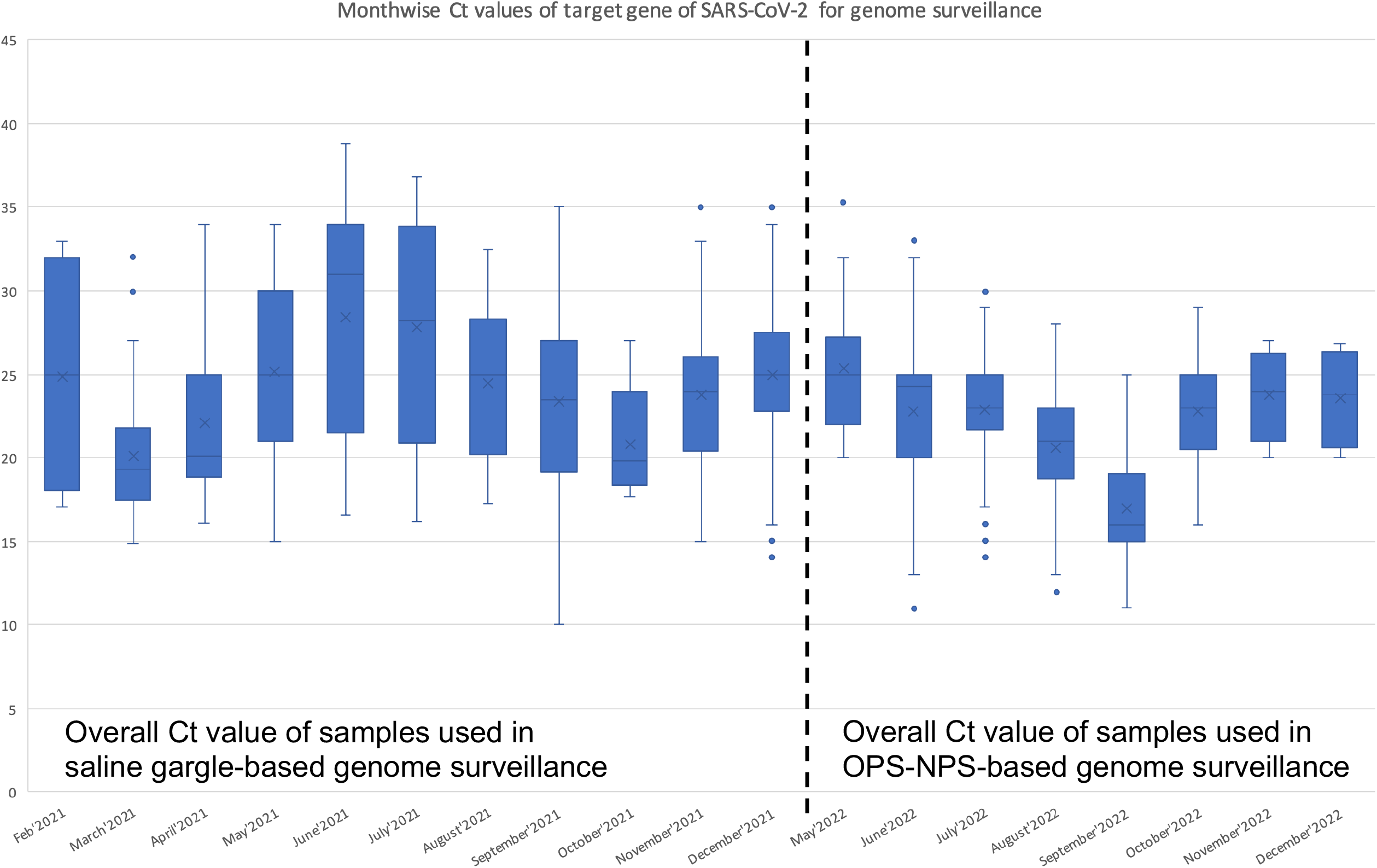
Monthwise distribution of Ct value for SARS-CoV-2 samples used in saline gargle and OPS-NPS-based genome surveillance.

### 2.7 Bioinformatic analysis

#### 2.7.1 Sequencing data processing

The live base-calling was performed using the Guppyv22.10.7 base-calling algorithm integrated into the MinION Mk1C sequencer [18]. The processed FASTQ reads from the MinION sequencer were analysed using a graphical user interface bioinformatics platform COMMANDER developed by Genotypic Technology Pvt. Ltd [19]. The FASTA sequence and the variant call performed by the COMMANDER pipeline were further confirmed by the web-based Pangolin COVID-19 Lineage Assigner [20]. The sequencing quality control criteria of >70% horizontal genome covered and >100X depth coverage were applied. The metadata of 3002 samples, including the FASTA sequence, was submitted to the Global initiative on sharing Avian Influenza Data (GISAID) [21] and the Indian biological data centre (IBDC) [22].

#### 2.7.2 Mutation Mapping

The mutation mapping of SARS-CoV-2 variants was done using the Outbreak.info web server [23]. Outbreak.info server calculates the prevalence of mutations as a ratio of the number of sequences carrying a given set of mutations on a given day at a specific place (or all locations) (x) divided by the total number of sequences on that day in that location (n). A 95 % Confidence interval was calculated using Jeffrey’s Interval: 2.5 quantiles of (x+0.5, n-x+0.5) to 97.5 quantiles of (x+0.5, n-x+0.5). GISAID assigns PANGO lineage for each unique sequence. The Pango nomenclature system’s classification is subject to changes as new lineages are uploaded to the GISAID database. While fundamental sequencing data remains the same in these cases, reporting for particular lineages may vary if analysed during a different period. The criteria of mutations with >75% prevalence in at least one lineage were applied.

## 3. RESULTS AND DISCUSSION

The two-year-long study used oxford nanopore technologies (ONT) based next-generation sequencing (NGS) platform. This study identified 104 different SARS-CoV-2 pango-lineages classified into 19 clades (**Figure 3)**. The detailed metadata of 3002 samples has been uploaded on the GISAID portal, and the accession numbers of the published sequences are provided in the supplementary file.

**Figure 3:**
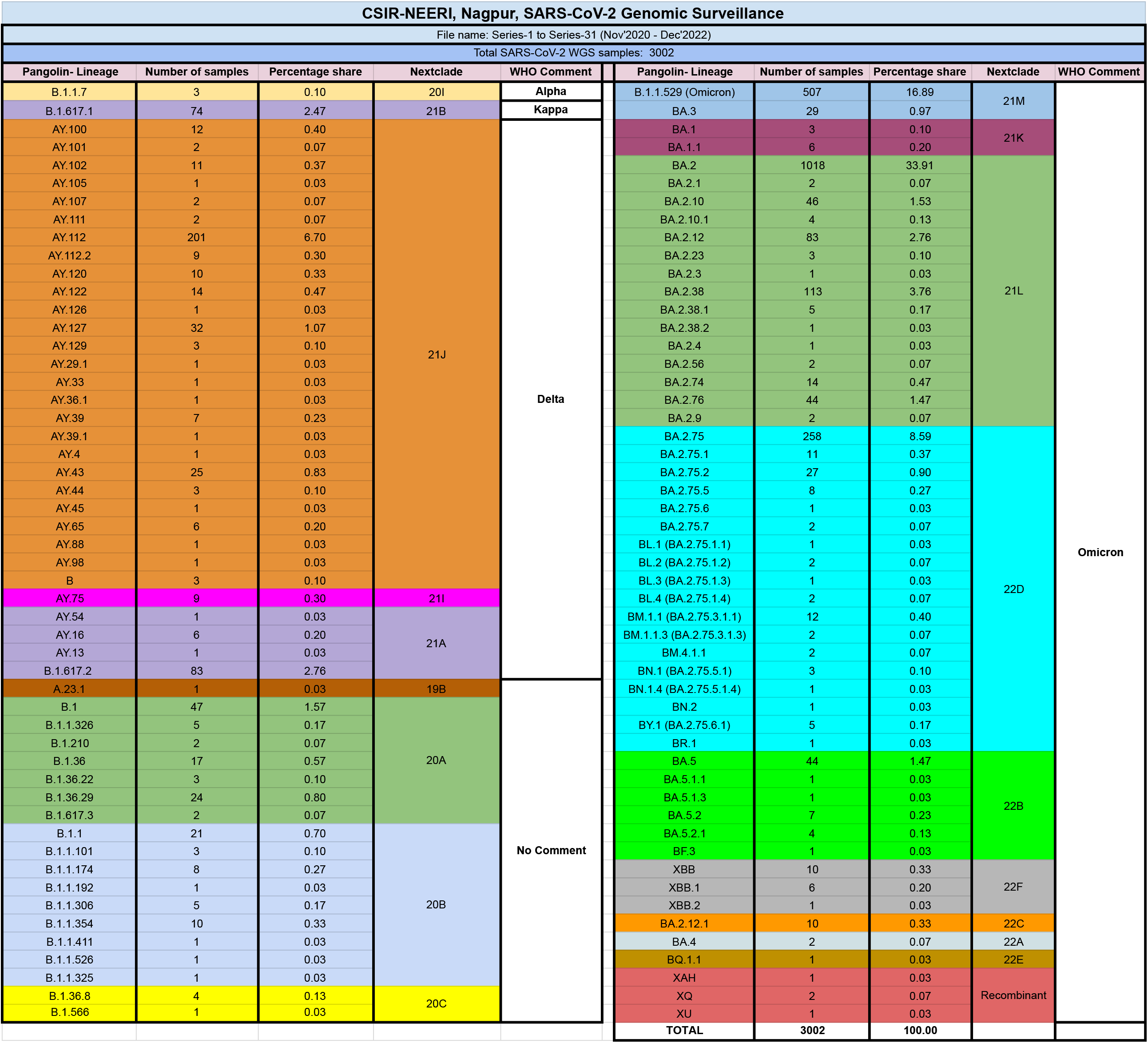
Summary of pango-lineages and their corresponding clades found in the SARS-CoV-2 genome surveillance.

The study revealed a diverse variant profile of 104 pango-lineages, which included 13 major variants with >1% prevalence among the total variants like B.1.1.529 (Omicron), (16.89%), BA.2 (33.91%), BA.2.75 (8.59%), AY.112 (6.70%), BA.2.38 (3.76%), B.1.617.2 (2.76%), BA.2.12 (2.76%), B.1.617.1 (2.47%), B.1 (1.57%), BA.2.10 (1.53%), BA.5 (1.47%), BA.2.76 (1.47%), and AY.127 (1.07%) (**Figure 4)**. As per the variant prevalence data retrieved from the open-sourced covlineages.org [24], the prevalence of variants in other countries is as follows, B.1.1.529 (Omicron) was prevalent in the United States of America at 48.0% and Germany at 11.0%. BA.2 was prominent in the United Kingdom at 29.0%, Germany at 13.0%, the United States of America at 12.0%, Denmark at 12.0%, and France at 7.0%. BA.2.75, one of Omicron’s most widely circulating sublineages, was prevalent in India at 72.0%, the United States of America at 5.0%, Canada at 3.0%, Japan at 2.0%, and Australia at 2.0%. AY.112, a member of Delta, was majorly prevalent in India at 34.2%, Germany at 11.2%, and Norway at 5.9%. Variant BA.2.38 was prevalent in India at 59.0%, the United States of America at 12.0%, the United Kingdom at 7.0%, Australia at 3.0%, and France at 2.0%. B.1.617.2 showed prevalence in India at 23.0%, Turkey at 16.0%, the United States of America at 14.0%, Germany at 7.0%, and the United Kingdom at 7.0%. BA.2.12 was prevalent in the United States of America at 54.0%, the United Kingdom at 14.0%, Germany at 9.0%, France at 6.0%, and India at 3.0%. B.1.617.1 was prevalent in the United States of America at 46.0%, Turkey at 11.0%, the United Kingdom at 6.0%, Canada at 4.0%, and France at 3.0%. BA.2.10 had shown the prevalence in India at 32.0%, the United Kingdom at 27.0%, the United States of America at 10.0%, Japan at 9.0%, and New Zealand at 3.0%. BA.5, a subtype of Omicron, was prevalent in the United States of America at 36.0%, Germany at 12.0%, France at 9.0%.BA.2.76 was prevalent in India at 67.0%, United States of America at 10.0%, Australia at 3.0%, United Kingdom at 3.0%, Japan at 2.0%. The variant AY.127 was India at 21.8%, Norway at 8.3%, and the United Kingdom at 13.2%.

**Figure 4:**
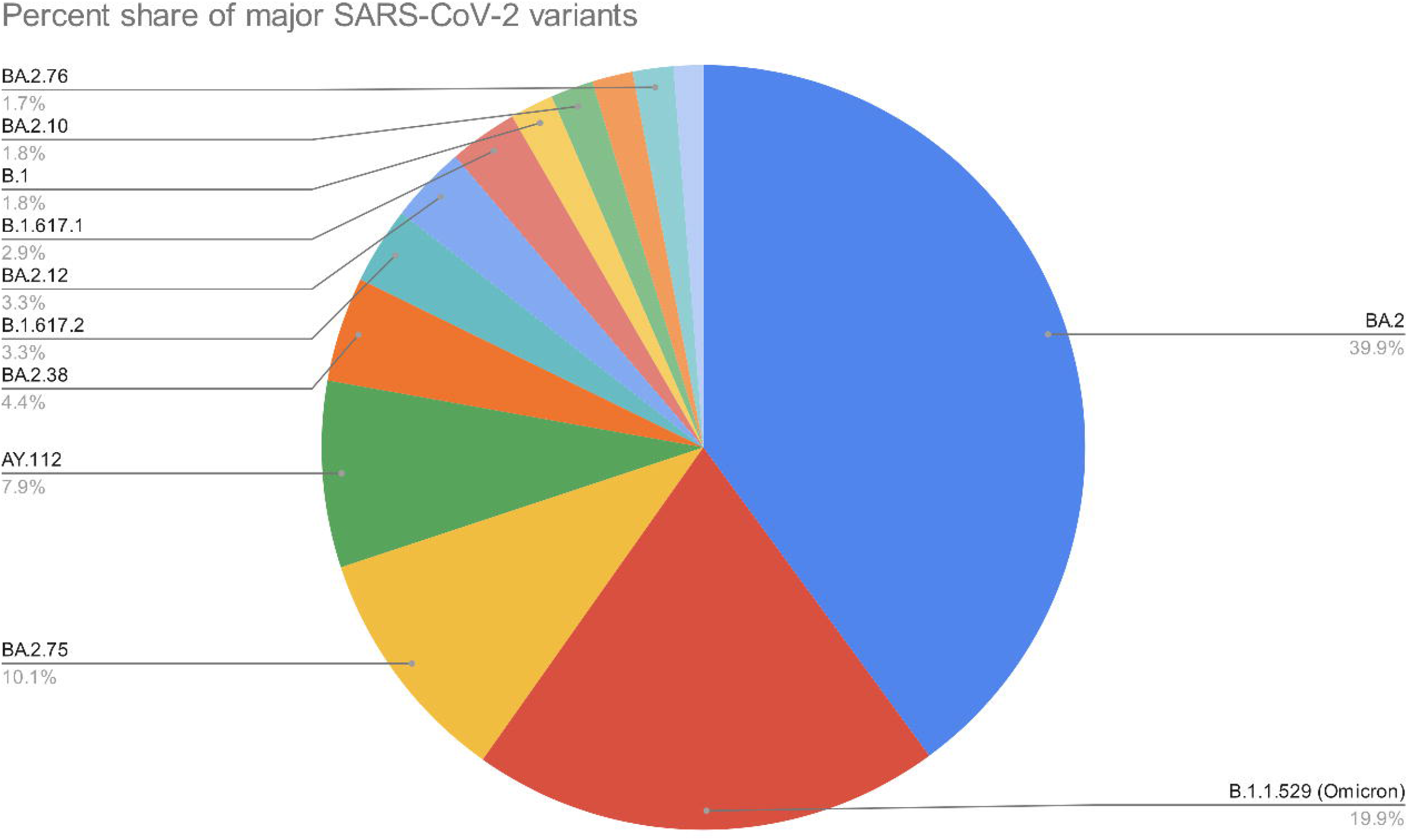
Pie-chart summarising major pango-lineages found in the SARS-CoV-2 genome surveillance.

In this study, the variants of the Omicron lineage have dominated the variant distribution post-December 2021; the omicron variants reported in this study belong to 10 different clades, as shown in **Figure 3**. The Omicron variant was first detected in Botswana and South Africa on 24 November 2021 [25] and was declared as a variant of concern (VOC) by the world health organisation (WHO) on 26 November 2021. Within one month of first reporting in Botswana, the B.1.1.529 (Omicron) variant was reported in our study from a sample collected on 21 December 2021. The sample was a 22-year-old female from Nagpur city in Maharashtra without international travel history. The prevalence of variants belonging to the Omicron lineage continued to proliferate and dominated the variant distribution afterwards. Other Omicron sub-variants, such as BA.1, BA.2, BA.3, BA.4, and BA.5, evolved from B.1.1.529 [26]. Two sub-variants of BA.5 have evolved, BQ.1 and BQ.1.1. The Omicron variant is reported to be an immune escape variant as it has caused multiple cases of infection in vaccinated individuals [27]. A study by Chakraborty et al. showed that the mutations D614G, E484A, N501Y, K417N, Y505H, and G496S improved S-glycoprotein accessibility to engage with the ACE2 receptor, which may contribute to enhancing the infectivity of Omicron variant [28].

### 3.1 FROM KAPPA TO OMICRON VIA DELTA

The genomic surveillance of SARS-CoV-2 started in November 2020, and lineage B.1 dominated the distribution with 100% prevalence in November and December 2020. Variant B.1.617.1, also known as the Kappa variant, started appearing in January 2021 and dominated the distribution till March 2021. The variants of Delta lineages B.1.617.2 and AY.112 also started appearing in February 2021. The first Omicron variant, B.1.1.529, appeared in December 2021. The month-wise dynamics of SARS-CoV-2 over two years clearly showed the complete transition of SARS-CoV-2 from delta to omicron variant in January of 2022. The detailed month-wise distribution, along with relative percentage share, is elaborated in **Figures 5a and 5b**

**Figure 5a:**
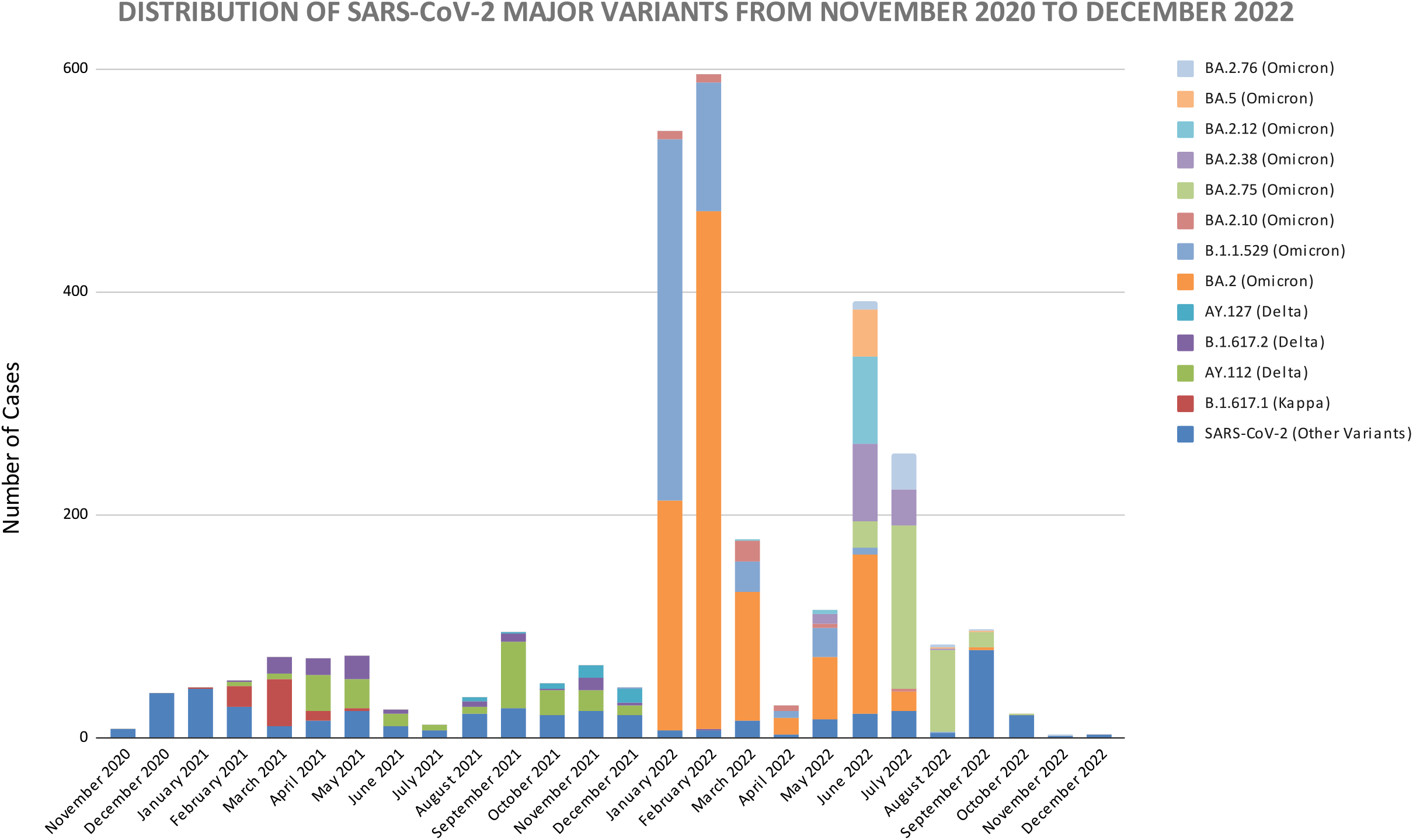
Monthly distribution of major pango-lineages found in the SARS-CoV-2 genome surveillance.

**Figure 5b:**
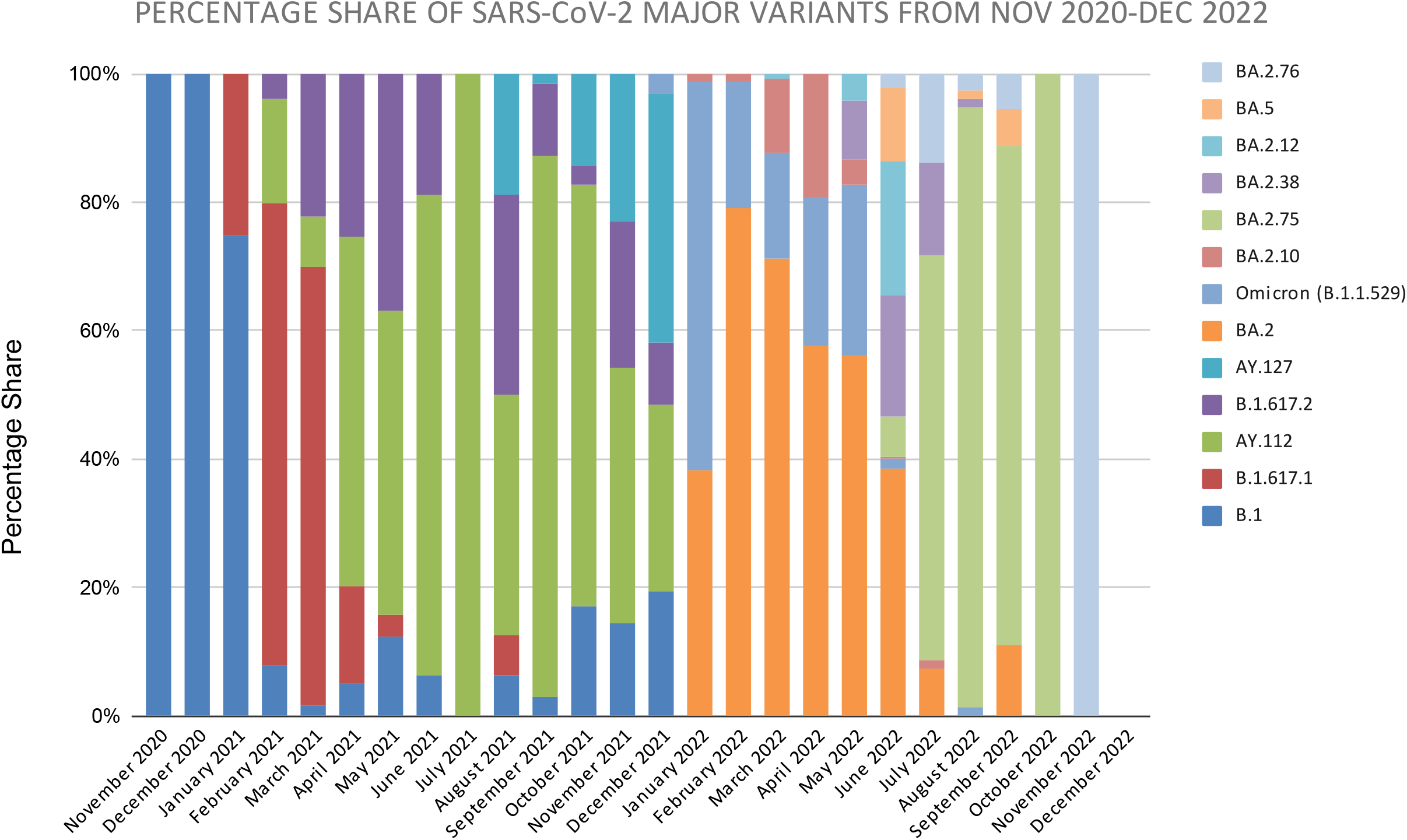
Percentage-wise monthly distribution of major pango-lineages found in the SARS-CoV-2 genome surveillance.

### 3.2 EPIDEMIOLOGICALLY SIGNIFICANT AND EMERGING SARS-CoV-2 LINEAGES

This study observed some epidemiologically significant variants from August 2022 to December 2022. The variants BM.1.1 (BA.2.75.3.1.1) and BM.1.1.3 (BA.2.75.3.1.3) belonging to the 22D clade were observed in September 2022; the BM.1.1.3 variant was majorly reported from India(20.1%), USA (14.8%), and United Kingdom (8.4%) [24]. Another important group of variants belonged to the clade 22F: XBB, XBB.1, and XBB.2. The XBB and its related variants were responsible for the increase in SARS-CoV-2 cases in the USA, Singapore, and Europe [29]. XBB.1.5 has become the most common variant in the USA since December 2022. Since February 2023, the XBB.1.16 has been India’s most dominating SARS-CoV-2 variant, as per the GISAID database [21]. XBB variants show high immune evasion and increased hACE2 binding due to an F486S amino acid substitution in the spike glycoprotein [30].

### 3.3 HOSPITALISATION AND DEATH CASES

The study has also evaluated the hospitalisation rate, reinfection, and death rate among Vaccinated and Unvaccinated patients. the vaccination data of 1636 patients were available, out of which 1232 were unvaccinated and 404 were vaccinated. In the unvaccinated group, 106 patients (8.6%) were hospitalised, 1029 (83.5%) were home-isolated, 13 (1%) were reinfection cases, and 10 (0.8%) patients died due to the SARS-CoV-2 infection. In the vaccinated patient group, 39 patients (9.6%) were hospitalised, 320 (79.2%) were home-isolated, 3 (0.7%) were reinfection cases, and no patient died due to the SARS-CoV-2 infection. The data indicate that among the vaccinated patient group, there was no death reported, and the incidences of reinfection cases were also lower in the vaccinated patient group (**Figure 6)**.

**Figure 6:**
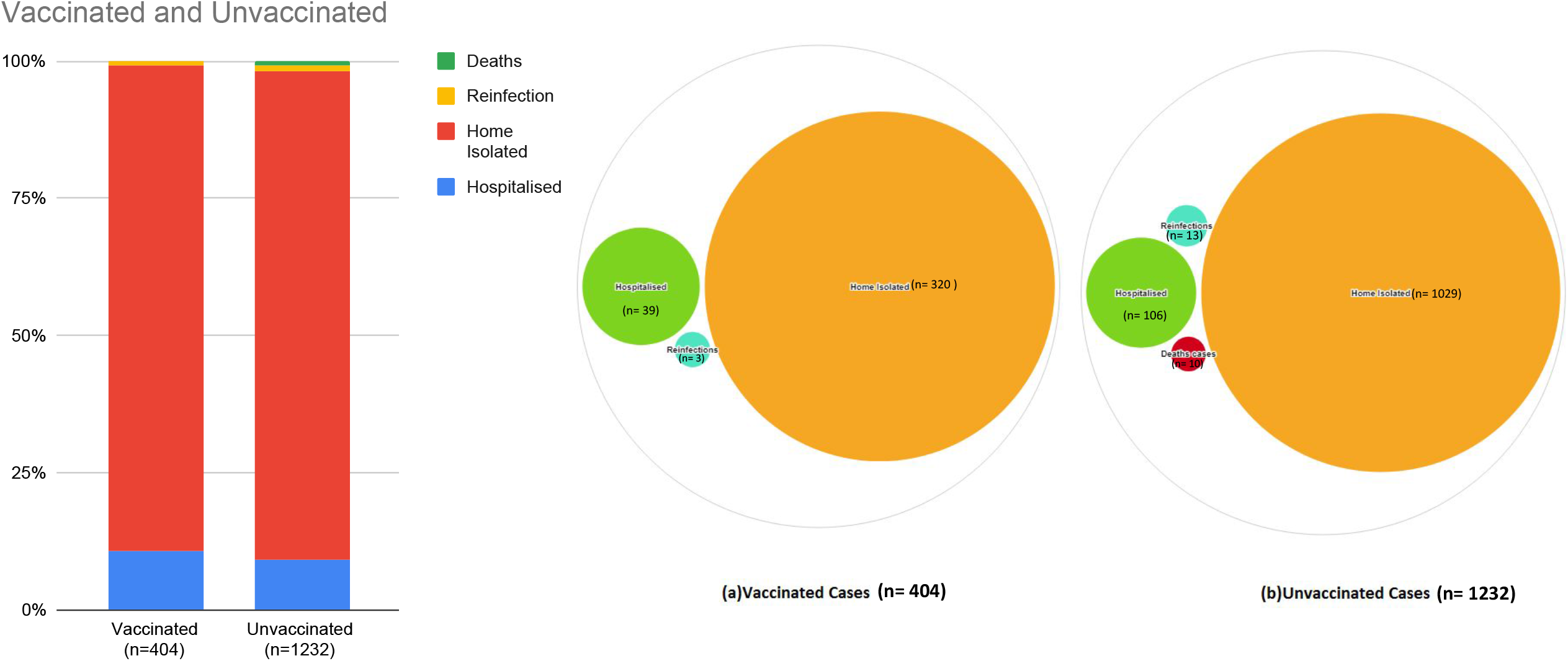
Summary of hospitalisation, home isolation, reinfection, and death rate among vaccinated and unvaccinated cases in the study.

### 3.4 VARIANTS PREVAILING IN THE VACCINATED POPULATION

Among the vaccinated patients, the vaccine type information was available for 122 patients, out of which 88 were vaccinated with Oxford-AstraZeneca vaccine (ChAdOx1), also known as Covishield, manufactured under licence by Serum Institute of India [31] and 33 were vaccinated with indigenously developed Covaxin (BBV152) [32]. One patient was vaccinated with the Russian Sputnik V vaccine [33]. The major SARS-CoV-2 variant prevailing among vaccinated patients who received Covishield or Covaxin was BA.2, followed by other variants of Omicron (**Figure 7)**. These study results indicate the immune escape properties of the Omicron variant. WHO, in its Weekly epidemiological update on COVID-19 issued on 14 December 2021, indicated that after the 15th week, the original two-dose regimen of the ChAdOx1 vaccine is ineffective against symptomatic illness caused by the Omicron variant. Andrews et al. (2022) also showed that the ChAdOx1 vaccine showed only 17.8% effectiveness against the Omicron variant of SARS-CoV-2 after 15-19 weeks post-second dose [34]. These findings may explain the sudden rise in the Omicron cases in India during December 2021. The vaccine effectiveness may have waned off and rendered ineffective in protecting the population against the omicron variant after 15-19 weeks post-vaccination.

**Figure 7:**
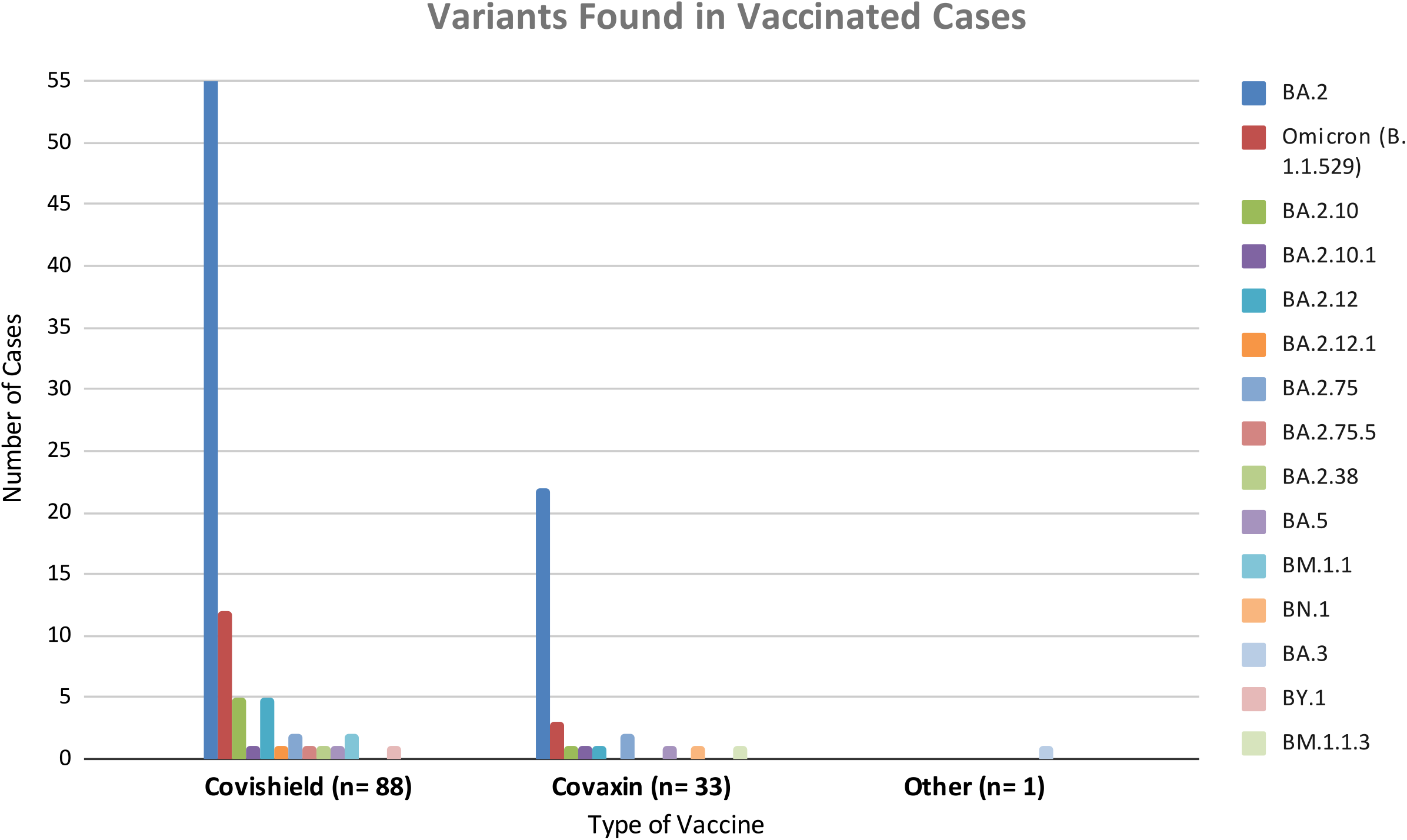
Summary of SARS-CoV-2 variants prevailed among vaccinated cases who received Covishield, Covaxin and Sputnik V.

### 3.5 MUTATION PROFILE OF THE VARIANTS FOUND IN THE STUDY

An increase in infectivity and immune escape of the SARS-CoV-2 virus results from mutations in the SARS-CoV-2 genome. We have analysed the mutation profiles of major variants in our study using the Outbreak.info web server [23] **(Figure 8a)**. The analysis showed seven mutations of interest (MOI) which include K417N, K417T, L452R, S477N, N501Y, P681H, P681R and one mutation of concern (MOC) E484K in the spike glycoprotein region **(Figure 8a)**. The K417N mutation was found to be >60% MOI prevalence among the GISAID sequences of B.1.1.529, BA.2, BA.2.10, BA.2.12, BA.2.38, BA.2.76, BA.2.75, and BA.5; while the Mutation K417T appeared with >60% MOI prevalence in BA.2.38. L452R mutation was observed among the AY.112, B.1.617.2, and AY.127 variants of delta lineage with >90% MOI prevalence. Interestingly, the L452R mutation was also noticed in the BA.5 variant of the Omicron lineage, recognised as a delta-like mutation [35]. Mutations S477N, N501Y and P681H showed 65-90% MOI prevalence in B.1.1.529, BA.2, BA.2.10, BA.2.12, BA.2.38, BA.2.76, BA.2.75, and BA.5. However mutation P681R appeared in the variants of delta lineages AY.112, B.1.617.2, and AY.127 with >90% MOI prevalence. The E484K mutation showed <0.1% MOC prevalence.

**Figure 8a:**
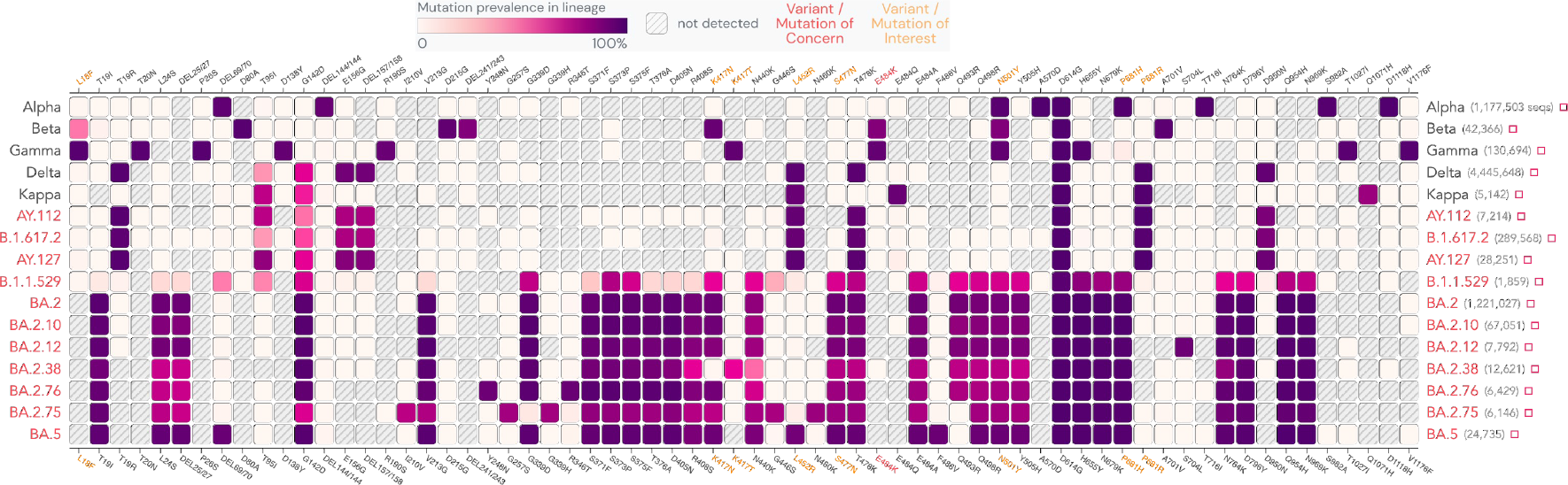
Mutation mapping of the Spike glycoprotein gene for the major SARS-CoV-2 lineages in the study.

**Figure 8b:**
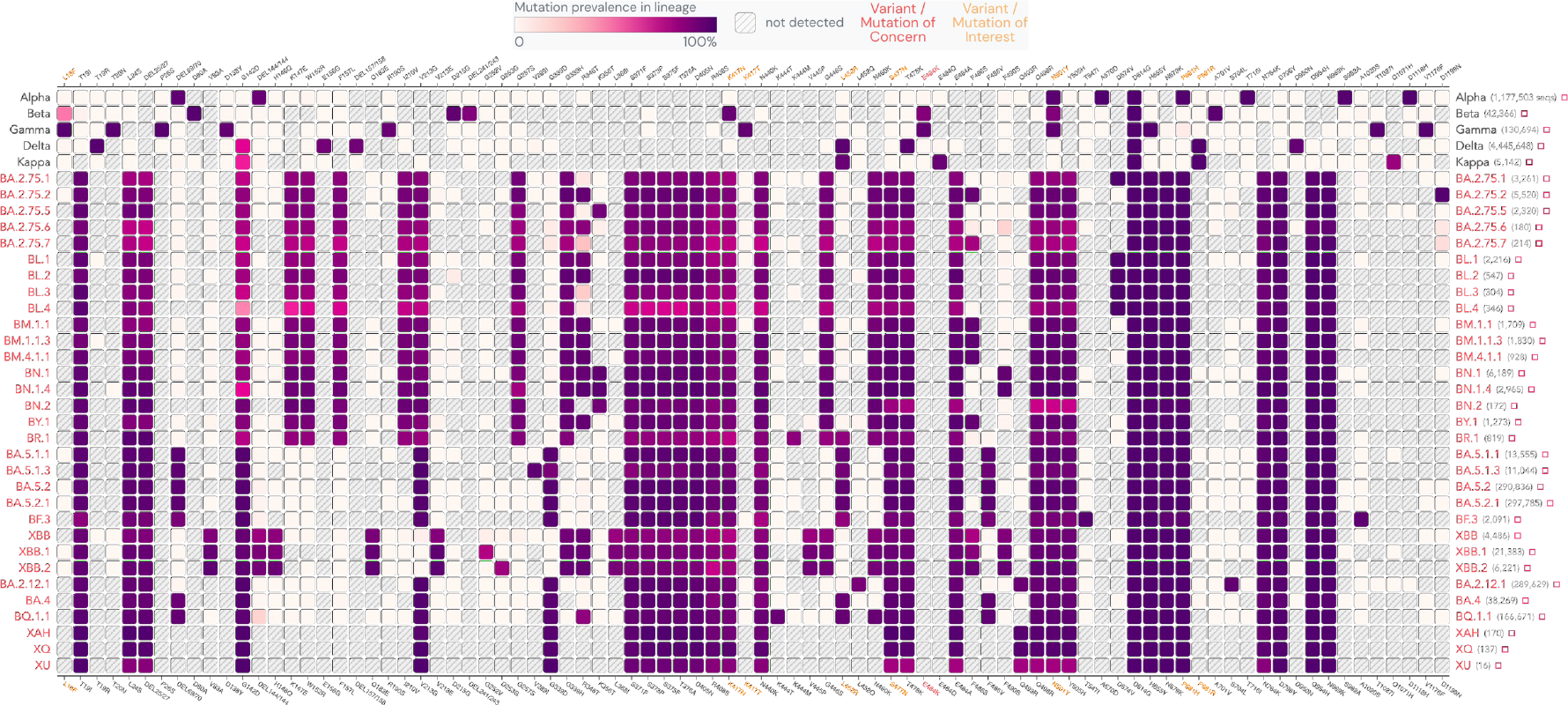
Mutation mapping of the Spike glycoprotein gene for the epidemiologically significant and emerging SARS-CoV-2 lineages in the study.

We have also analysed the mutation profiles of some epidemiologically significant variants found in the later phase of the study using the Outbreak.info web server **(Figure 8b)**. The analysis showed eight MOI, including L18F, K417N, K417T, L452R, S477N, N501Y, P681H, P681R, and MOC E484K in the spike glycoprotein region **(Figure 8b)**. The L18F mutation showed <0.1% MOI prevalence in BA.2.75.2, BA.5.1.1, BA.5.2, BA.5.2.1, XBB.1, and XBB.2. The mutation profile showed that all the variants isolated in the study retained the MOI K417N with 70-90% MOI prevalence. The K417T mutation showed <0.1% MOI prevalence in BA.2.75.21, BA.2.75.2, BA.2.75.6, BA.2.75.7, BL.1, BM.1.1, BF.3, BA.2.12.1, BA.4 and BQ.1.1. The L452R mutation showed 70-95% MOI prevalence among the BR.1, BA.5.1.1, BA.5.1.3, BA.5.2, BA.5.2.1, BF.3, BA.4, and BQ.1.1. The mutation profile showed that all the variants isolated in the study retained the MOI S477N and N501Y with 80-90% MOI prevalence. The mutation profile showed that all the variants isolated in the study retained the MOI P681H with 90-100% MOI prevalence. The P681R mutation showed <0.1% MOI prevalence in BN.1, BA.5.1.1, BA.5.2, BA.5.2.1, XBB.1, BA.2.12.1 and BA.4. The MOC E484K consistently remain undetected among the variants found in the later part of the study.

L18F substitution is one of the most prevalent mutations sequenced to date and is present in both the B.1.351 and P.1 lineages [36]. L18F is associated with escaping N-terminal domain binding antibodies [37]. K417N mutation had reduced neutralisation by Casirivimab (REGN10933) and Etesevimab monoclonal antibodies [38]. K417T mutations showed stronger binding with the hACE2 receptor contributing to increased pathogenicity of the variants possessing this mutation [39]. By December 2020 and February 2021, L452R occurred independently in multiple lineages worldwide, indicating that this amino acid substitution is most likely the consequence of viral adaptability due to increased immunity in the population [40]. The L452R mutation in RBD reduces the neutralising action of RBD-specific mAbs such as regdanvimab (CT-P59), etesevimab (LY-CoV016), and bamlanivimab (LY-CoV555) [41]. The S477N mutation is reported for a 37-fold increase in the RBD/ACE2 binding, thereby increasing the infectivity of variants having this mutation [42]. In vitro studies showed that the mutation N501Y resulted in a significant increase in ACE2 affinity imparted by a single RBD mutation [43]. Studies using pseudoviruses with N501Y RBD mutation revealed that the neutralising activity of plasma from vaccinated individuals decreased significantly [44]. The mutation P681H is reported to confer resistance against innate immune response [45]. According to a study, the P681R mutation might be connected to the increased frequency of non-synonymous mutations in SARS-CoV-2 variants found in Maharashtra during February 2021 [40]. The mutation P681R in the furin cleavage site led to increased basicity of the poly-basic stretch that might improve the transmissibility of the SARS-CoV-2 virus [45]. The mutation E484K was discovered to be one of the most prevalent mutations to evade monoclonal antibodies and natural class 2 polyclonal antibody response [46,47]. E484K is among the most common RBD mutations reported in in-vitro studies and sequences on the GISAD database.

## Data Availability

All data produced in the present study are available upon reasonable request to the authors

## Data Availability

All data produced in the present study are available upon reasonable request to the authors

## Data Availability

All data produced in the present study are available upon reasonable request to the authors

## FIGURE LEGEND

**Supplementary file:** Line listing of GISAID submission with accession number for 3002 SARS-CoV-2 genome sequences from the study

## ACKNOWLEDGMENTS

This work was supported and funded by CSIR-NEERI, Nagpur. It is certified that the manuscript has been checked for plagiarism by the institute knowledge resource centre through iThenticate (anti-plagiarism software) KRC No. CSIR-NEERI/KRC/2023/MARCH/EVC/2.

## REFERENCES

1. Harvey, W.T., Carabelli, A.M., Jackson, B. et al. SARS-CoV-2 variants, spike mutations, and immune escape. Nat Rev Microbiol 19, 409–424 (2021). https://doi.org/10.1038/s41579-021-00573-0

2. Mengist, H. M., Kombe Kombe, A. J., Mekonnen, D., Abebaw, A., Getachew, M., &amp; Jin, T. (2021). Mutations of SARS-COV-2 spike protein: Implications on immune evasion and vaccine-induced immunity. Seminars in Immunology, 55, 101533. https://doi.org/10.1016/j.smim.2021.101533

3. Genome Sequencing by INSACOG shows variants of concern and a Novel variant in India. (2021, March 21). pib.gov.in. Retrieved March 13, 2023, from https://pib.gov.in/PressReleaseIframePage.aspx?PRID=1707177

4. Kay, C., & Pandya, D. (2021, December 29). How Errors, Inaction Sent a Deadly Covid Variant Around the World. Bloomberg.com. https://bloomberg.com/news/features/2021-12-29/how-delta-variant-spread-in-india-deadly-errors-inaction-covid-crisis

5. Koshy, J. (2021, October 3). The story of how a mystifying novel coronavirus variant, Delta, has India and the globe in its grip. https://www.thehindu.com/sci-tech/health/the-story-of-how-a-mystifying-novel-coronavirus-variant-delta-has-india-and-the-globe-in-its-grip/article36772942.ece

6. Provisional Population Totals, Census of India 2011 Cities Having Population 1 Lakh and above Provisional Population Totals, Census of India 2011 Cities Having Population 1 Lakh and Above.; 2011.https://www.census2011.co.in/city.php

7. Khairnar, K., & Tomar, S. S. (2023). Utility of non-invasive Saline Gargle SARS-CoV-2 genome surveillance in post-pandemic scenario and limitations of wastewater surveillance. MedRxiv (Cold Spring Harbor Laboratory). https://doi.org/10.1101/2023.02.16.23286031

8. IHIP. Integrated Health Information Platform Integrated Disease Surveillance Programme Ministry of Health and Family Welfare, Government of India @ ihip.nhp.gov.in. https://ihip.nhp.gov.in/idsp/#!/.

9. ICMR. ICMR Public Dashboard @ influenza.icmr.org.in https://influenza.icmr.org.in/public_dashboard/ Accessed on March 22, 2023.

10. WHO. Global Epidemiological Surveillance Standards for Influenza.; 2013. https://www.who.int/publications/i/item/9789241506601 Accessed on March 22, 2023

11. Director CSIR-NEERI. (2021b, August 9). Saline Gargle RT PCR Innovation by CSIR NEERI [Video]. YouTube. https://www.youtube.com/watch?v=1OPZFlF4GpQ

12. Himedia. Insta NX ® Viral RNA Purification Kit.; 2022. https://www.himedialabs.com/in/mbin013-insta-nx-viral-rna-purification-kit.html.

13. Hi-PCR® COVID-19 triplex probe PCR kit. HiMedia Leading BioSciences Company. Retrieved March 22, 2023, from https://www.himedialabs.com/in/hi-pcrr-covid-19-triplex-probe-pcr-kit.html

14. RT PCR KIT: COVID-19 one-step rt-PCR kit. Meril Life. Retrieved March 22, 2023, from https://www.merillife.com/assets/pdfs/medical-devices/covid-19-one-step-rt-pcr-kit-1608096936pdf.pdf

15. WHO. End-to-end integration of SARS-CoV-2 and influenza sentinel surveillance. 2022 https://www.who.int/publications/i/item/WHO-2019-nCoV-Integrated_sentinel_surveillance-2022.1

16. Takara bio inc. RR037A For Research Use PrimeScript TM RT Reagent Kit (Perfect Real Time) Product Manual. https://www.takarabio.com/documents/UserManual/RR037A_e.v2008Da.pdf.

17. Oxford Nanopore Technologies. PCR Tiling of SARS-CoV-2 Virus with Rapid Barcoding and Midnight RT PCR Expansion (SQK-RBK110.96 and EXPMRT001). Vol 3.; 2021. https://nanoporetech.com/resource-centre/knowledge Accessed on March 22, 2023

18. Wick RR, Judd LM, Holt KE. Performance of neural network base-calling tools for Oxford Nanopore sequencing. Genome Biol. 2019:1–10. https://doi.org/10.1186/s13059-019-1727-y

19. Commander, a command line free GUI-based sequencing data analysis tool developed by Genotypic Technology, Bangalore India https://www.genotypic.co.in/commander/. Accessed on March 22, 2023

20. Toole ÁO, Scher E, Underwood A, et al. Assignment of epidemiological lineages in an emerging pandemic using the pangolin tool. 2021;7(2):1–9. https://doi.org/10.1093/ve/veab064

21. gisaid.org. 2022. https://gisaid.org/submission-tracker-global/ (accessed March 22, 2023)

22. INSACOG - INDA-CA. INDA. Retrieved March 22, 2023, from https://inda.rcb.ac.in/insacog/indexpage

23. Outbreak.info SARS-COV-2 data explorer. outbreak.info. Retrieved March 22, 2023, from https://outbreak.info/

24. Lineages.org. Cov. Retrieved March 22, 2023, from https://cov-lineages.org/

25. Mallapaty, S. (2022). Where did Omicron come from? Three key theories. Nature, 602(7895), 26–28. https://doi.org/10.1038/d41586-022-00215-2

26. Shrestha, L. B., Foster, C., Rawlinson, W., Tedla, N., &amp; Bull, R. A. (2022). Evolution of the sarslJcovlJ2 omicron variants BA.1 to Ba.5: Implications for immune escape and transmission. Reviews in Medical Virology, 32(5). https://doi.org/10.1002/rmv.2381

27. Cao, Y. R., Wang, J., Jian, F., Xiao, T, et al. Omicron escapes the majority of existing SARS-CoV-2 neutralizing antibodies. Nature, 602(7898), 657–663. https://doi.org/10.1038/s41586-021-04385-3

28. Chakraborty, C., Bhattacharya, M., Sharma, A., & Mallik, B. (2022). Omicron (B.1.1.529) - A new heavily mutated variant: Mapped location and probable properties of its mutations with an emphasis on S-glycoprotein. International Journal of Biological Macromolecules, 219, 980–997. https://doi.org/10.1016/j.ijbiomac.2022.07.254

29. Callaway, E. (2023, January 9). Coronavirus variant XBB.1.5 rises in the United States - is it a global threat? Nature News. Retrieved March 22, 2023, from https://www.nature.com/articles/d41586-023-00014-3

30. Salsekar, L., Rahangdale, S., Tamboli, E., & Khairnar, K. (2023). Unique amino acid substitution in RBD region of SARS-COV-2 omicron XAY.2. https://doi.org/10.1101/2023.01.09.523246

31. World Health Organization: WHO. (2022, June 13). The Oxford/AstraZeneca (ChAdOx1-S [recombinant] vaccine) COVID-19 vaccine: what you need to know. https://www.who.int/news-room/feature-stories/detail/the-oxford-astrazeneca-covid-19-vaccine-what-you-need-to-know

32. World Health Organization: WHO. (2022a, June 10). The Bharat Biotech BBV152 COVAXIN vaccine against COVID-19: What you need to know. https://www.who.int/news-room/feature-stories/detail/the-bharat-biotech-bbv152-covaxin-vaccine-against-covid-19-what-you-need-to-know

33. About Sputnik V. Official Website Vaccine Against COVID-19 Sputnik V. Retrieved March 22, 2023, from https://sputnikvaccine.com/about-vaccine/

34. Andrews, N., Stowe, J., Kirsebom, F. C. M., Toffa, S. E., Rickeard, T., Gallagher, E., Gower, C., Kall, M., Groves, N., O’Connell, A., Simons, D. S., Blomquist, P., Zaidi, A. A., Nash, S., Aziz, N. A., Thelwall, S., Dabrera, G., Myers, R. M., Amirthalingam, G. Bernal, J. L. (2022). Covid-19 Vaccine Effectiveness against the Omicron (B.1.1.529) Variant. The New England Journal of Medicine, 386(16), 1532–1546. https://doi.org/10.1056/nejmoa2119451

35. Islam MR, Shahriar M, Bhuiyan MA. The latest Omicron BA.4 and BA.5 lineages are frowning toward COVID-19 preventive measures: A threat to global public health. Health Sci Rep. 2022 Oct 13;5(6):e884. https://doi.org/10.1002/hsr2.884

36. Harvey, W.T., Carabelli, A.M., Jackson, B. et al. SARS-CoV-2 variants, spike mutations and immune escape. Nat Rev Microbiol 19, 409–424 (2021). https://doi.org/10.1038/s41579-021-00573-0

37. McCallum, Matthew, De Marco, Anna, Lempp Florian A., Tortorici, M. A., Pinto, Dora, Walls, Alexandra C., Beltramello, Martina et al. “N-terminal domain antigenic mapping reveals a site of vulnerability for SARS-CoV-2.” Cell 184, no. 9 (2021): 2332-2347.e16. Accessed March 21, 2023. https://doi.org/10.1016/j.cell.2021.03.028.

38. Tada, T., Zhou, H., Dcosta, B. M., Samanovic, M. I., Chivukula, V., Herati, R. S., Hubbard, S. R., Mulligan, M. J., &amp; Landau, N. R. (2021). Increased resistance of SARS-COV-2 omicron variant to neutralization by vaccine-elicited and therapeutic antibodies. https://doi.org/10.1101/2021.12.28.474369

39. Khan, A., Zia, T., Suleman, M., Khan, T., Ali, S. S., Abbasi, A. A., Mohammad, A., & Wei, D. (2021). Higher infectivity of the SARS-CoV-2 new variants is associated with K417N/T, E484K, and N501Y mutants: An insight from structural data. J Cell Physiol, 236, 7045–7057. https://doi.org/10.1002/jcp.30367

40. Cherian, S., Potdar, V., Jadhav, S., Yadav, P., Gupta, N., Das, M., Rakshit, P., Singh, S., Abraham, P., &amp; Panda, S. (2021). Convergent evolution of SARS-COV-2 spike mutations, L452R, E484Q and P681R, in the second wave of covid-19 in Maharashtra, India. https://doi.org/10.1101/2021.04.22.440932

41. McCallum, M., Bassi, J., Marco, A. D., Chen, A., Walls, A. C., Iulio, J. D., Tortorici, M. A., Navarro, M.-J., Silacci-Fregni, C., Saliba, C., Agostini, M., Pinto, D., Culap, K., Bianchi, S., Jaconi, S., Cameroni, E., Bowen, J. E., Tilles, S. W., Pizzuto, M. S., … Veesler, D. (2021). SARS-COV-2 immune evasion by variant B.1.427/b.1.429. https://doi.org/10.1101/2021.03.31.437925

42. Dejnirattisai, W., Huo, J., Zhou, D., Zahradník, J., Supasa, P., Liu, C., Duyvesteyn, H. M. E., Ginn, H. M., Mentzer, A. J., Tuekprakhon, A., Nutalai, R., Wang, B., Dijokaite, A., Khan, S., Avinoam, O., Bahar, M., Skelly, D., Adele, S., Johnson, S. A., … Screaton, G. R. (2021). Omicron-B.1.1.529 leads to widespread escape from neutralizing antibody responses. https://doi.org/10.1101/2021.12.03.471045

43. Starr, T. N. et al. Deep mutational scanning of SARS-CoV-2 receptor binding domain reveals constraints on folding and ACE2 binding. Cell 182, 1295–1310.e1220 (2020). https://doi.org/10.1016/j.cell.2020.08.012.

44. Xie, X. et al. Neutralization of N501Y mutant SARS-CoV-2 by BNT162b2 vaccine-elicited sera. Preprint at bioRxiv https://doi.org/10.1101/2021.01.07.425740

45. Lista, M. J., Winstone, H., Wilson, H. D., Dyer, A., Pickering, S., Galao, R. P., De Lorenzo, G., Cowton, V. M., Furnon, W., Suarez, N., Orton, R., Palmarini, M., Patel, A. H., Snell, L., Nebbia, G., Swanson, C., & Neil, S. J. D. (2022). The P681H Mutation in the Spike Glycoprotein of the Alpha Variant of SARS-CoV-2 Escapes IFITM Restriction and Is Necessary for Type I Interferon Resistance. Journal of virology, 96(23), e0125022. https://doi.org/10.1128/jvi.01250-22

46. Yiska Weisblum, Fabian Schmidt, Fengwen Zhang, Justin DaSilva, Daniel Poston et. al. (2020) Escape from neutralizing antibodies by SARS-CoV-2 spike protein variants eLife 9:e61312 https://doi.org/10.7554/eLife.61312

47. Wang, Z., Schmidt, F., Weisblum, Y. et al. mRNAvaccine-elicited antibodies to SARS-CoV-2 and circulating variants. Nature 592, 616–622 (2021). https://doi.org/10.1038/s41586-021-03324-6

